# Point-of-care lung ultrasonography for early identification of mild COVID-19: a prospective cohort of outpatients in a Swiss screening center

**DOI:** 10.1101/2021.03.23.21254150

**Authors:** Siméon Schaad, Thomas Brahier, Mary-Anne Hartley, Jean-Baptiste Cordonnier, Luca Bosso, Tanguy Espejo, Olivier Pantet, Olivier Hugli, Pierre-Nicolas Carron, Jean-Yves Meuwly, Noémie Boillat-Blanco

## Abstract

**Background:** Early identification of SARS-CoV-2 infection is important to guide quarantine and reduce transmission. This study evaluates the diagnostic performance of lung ultrasound (LUS), an affordable, consumable-free point-of-care tool, for COVID-19 screening.

**Methods:** This prospective observational cohort included adults presenting with cough and/or dyspnea at a SARS-CoV-2 screening center of Lausanne University Hospital between March 31^st^ and May 8^th^, 2020. Investigators recorded standardized LUS images and videos in 10 lung zones per subject. Two blinded independent experts reviewed LUS recording and classified abnormal findings according to pre-specified criteria to investigate their predictive value to diagnose SARS-CoV-2 infection according to PCR on nasopharyngeal swabs (COVID^pos^ vs COVID^neg^). We finally combined LUS and clinical findings to derive a multivariate logistic regression diagnostic score.

**Results:** Of 134 included patients, 23% (n=30/134) were COVID^pos^ and 77% (n=103/134) were COVID^neg^; 85%, (n=114/134) cases were previously healthy healthcare workers presenting within 2 to 5 days of symptom onset (IQR). Abnormal LUS findings were significantly more frequent in COVID^pos^ compared to COVID^neg^ (45% versus 26%, p=0.045) and mostly consisted of focal pathologic B-lines. Combining LUS findings in a multivariate logistic regression score had an area under the receiver-operating curve of 63.9% to detect COVID-19, but improved to 84.5% with the addition of clinical features

**Conclusions:** COVID^pos^ patients are significantly more likely to have lung pathology by LUS. Our findings have potential diagnostic value for COVID-19 at the point of care. Combination of clinical and LUS features showed promising results, which need confirmation in a larger study population.

**What is already known on the subject:**

- Lung ultrasonography (LUS) is a consumable-free, easy-to-use, portable, non-radiating and non-invasive screening tool that can be performed at the bedside: its diagnostic performance for pneumonia has been established.
- Recent studies conducted in emergency department showed a correlation between LUS findings and COVID-19 diagnosis.

**What the study ads:**

- This is the first study assessing the diagnostic performance of LUS for COVID-19 in outpatients with mild acute respiratory tract infection.
- Mild COVID-19 patients are more likely to have lung pathology by LUS compared with COVID-19 negative.
- Combination of clinical and LUS features showed promising results with a potential diagnostic value for COVID-19 at the point of care.

## Introduction

A year into the pandemic, Coronavirus Disease (COVID-19) remains a constant threat, overburdening the healthcare system. Current molecular diagnostic tests such as PCR and rapid antigen/antibody tests rely on consumables, which are vulnerable to shortages and saturation during exponential demand. The use of lung imaging as a diagnostic tool for COVID-19 has shown promises. Chest CT has a good sensitivity for patients triaged in emergency departments [1,2] and has even been able to detect pathology in asymptomatic cases, suggesting its potential as an early screening test in specific populations [3–5]. However, CT and even X-rays expose patients to ionizing radiation, are costly, and often not available in decentralized screening sites. Lung ultrasonography (LUS) is an alternative, consumable-free, easy-to-use, portable, non-radiating and non-invasive screening tool that can be performed at the bedside, with simple disinfection between patients and only a negligible cost of ultrasound gel as a consumable. It would allow immediate identification of infected patients at the point-of-care and be invaluable to the sustainable control of the pandemic. Its diagnostic performance for pneumonia has been established using chest CT as a gold standard [6]. For COVID-19, recent studies conducted in emergency departments showed several LUS patterns ranging from mild interstitial infiltrate, to lung consolidation, which correlated with disease progression and outcome [7,8]. However, these studies included mostly severe patients in emergency departments or intensive care units, which may lead to overoptimistic diagnostic performance of LUS due to a spectrum effect [9]. To our knowledge, no studies have described LUS findings in subjects with mild COVID-19. This study aims to compare LUS characteristics between SARS-CoV-2 PCR-confirmed (COVID^pos^) and PCR-negative (COVID^neg^) patients in a screening center and explore LUS performance for identification of COVID-19 outpatients.

## Methods

### Study design, setting and population

This prospective cohort study recruited consecutive outpatients at the COVID-19 screening center in Lausanne University Hospital, Switzerland (CHUV) between March 31^st^ and May 8^th^ 2020. All adults (age ≥ 18 years) presenting at the center with cough and/or dyspnea and who fulfilled eligibility criteria for nasopharyngeal SARS-CoV-2 real time (Rt-) PCR according to the State recommendations at the time of the study were eligible. These State criteria were the presence of symptoms suggestive of COVID in a health worker or a subject with at least one vulnerability criterion, *i*.*e*. age ≥ 65 years old or having at least one comorbidity (obesity, diabetes, active cancer, chronic cardiovascular, pulmonary, liver, renal or inflammatory disease). Exclusion criteria were uninterpretable Rt-PCR results or absence of LUS recording. Written informed consent was obtained from all participants.

To ensure that LUS abnormal findings would be specific of a respiratory tract infection, we included a control group of healthy volunteers, matched for age (± 5 years), sex, and smoking status with COVID^pos^ patients (Supplementary Table 1). These volunteers were asymptomatic during the previous 15 days (absence of odynophagia, cough, dyspnea, runny nose, fever, loss of smell or taste) and did not have a documented SARS-CoV-2 infection.

At inclusion, demographics, comorbidities, symptoms (including duration), and vital signs were collected using a standardized electronic case report form in REDCap® (Research Electronic Data Capture). Patients were subsequently classified as either COVID^pos^ or COVID^neg^ according to the SARS-CoV-2 RT-PCR results (at inclusion or at any time during the 30-day follow-up if the test was repeated for the same clinical episode). We assessed 30-day outcome by phone using a standardized interview (persistence of symptoms, secondary medical consultation, hospital admission, death).

The study was approved by the Swiss Ethics Committee of the canton of Vaud (CER-VD 2019-02283).

### Patient and public involvement

Subjects were not involved in the design or conduct of this study.

### Sample size

The minimum sample size required for this study was 100 patients with a clinical suspicion of COVID. It was calculated using a COVID prevalence of 20% and an estimated sensitivity of LUS to identify COVID^pos^ at 80% This sample size guarantees a power of 80% with a false discovery rate of 5% [10].

### Lung ultrasonography

Three medical students trained in LUS performed image acquisitions in the triage site. The first 10 acquisitions were done under direct supervision of an experienced board-certified expert (OP) who verified the quality of recorded images. Acquisition was standardized according to the “10-zone method” [11,12], consisting of five zones per hemithorax. Two images (sagittal and transverse) and 5 second videos were systematically recorded in every zone with a Butterfly IQ™ personal US system (Butterfly, Guiford, CT, USA), using the lung preset. The LUS probe and the electronic tablet were disinfected with an alcohol-based solution between each patient to avoid nosocomial spread [13].

For interpretation of LUS pathology, a physician experienced in LUS (TB) and an expert radiologist (JYM), blinded to patients’ diagnoses, independently filled a standardized report form as previously described [8]. Discordance between the two readers were adjudicated by a third expert (OP). The abnormal images were summed up in a LUS score for each patient, as previously described [8,14,15].

### Statistical analyses

Differences between COVID^pos^ and COVID^neg^ patients for all collected demographic and clinical features as well as LUS findings and LUS score were evaluated by Mann Whitney or chi-squared test, as appropriate. A bilateral p value <0.05 was considered as indicative of statistical significance. A multivariate logistic regression was built from 22, 15, 10 and 8 features using recursive feature elimination (RFE), originally including the following:

1. **LUS findings (n=10)**
  - Number of pathological zones for each of the five patterns (normal, pathological B lines, confluent B lines, pleural thickening, consolidation) (n=5)
  - A dichotomized variable for the presence/absence of the above four pathological patterns detected (n=4)
  - Binary variables for the presence of multifocal disease (n=1)
2. **Symptoms at presentation (n=8)**
  - Binary variables for the presence of cough, sputum, dyspnea, fever, anosmia, rhinorrhea, myalgia, and diarrhea
3. **Vital signs (n=3)**
  - Continuous variables for temperature, oxygen saturation, and respiratory rate
4. **Epidemiological history (n=1)**
  - Binary variable for a history of known unprotected contact with a COVID-19 case Feature coefficients are presented, as well as their importance in ranked order from RFE. Performance at several stages of the RFE are reported, using the top 22, 15, 10 and 8 features. Models using just LUS or just clinical findings were also built.

Diagnostic performance is reported as sensitivity, specificity, positive and negative predictive values (PPV, NPV), positive and negative likelihood ratios (LR+, LR-) and area under the receiver-operator curve (AUC). Due to the dataset size, we report findings on the entire dataset. A diagnostic score was derived from the summed coefficients, normalized within a range from −6 (COVID^pos^ highly unlikely) to +4 (COVID^pos^ highly likely) and the number of patients in each class are presented for each value of the score. The optimal cut-point was chosen using Youden index [16].

The kappa coefficient was calculated to measure the inter-rater agreement between the two LUS readers. R Core Team (2019) statistical software and python 3.0 with the sklearn library was used for analyses. Similar analyses were attempted on the outcome at 30-day follow up but impossible due to the limited sample size.

The reporting of our results followed the STARD guidelines.

## Results

### Demographics and clinical presentation

A total of 141 patients met inclusion criteria and were enrolled into the study; eight (5%) were later excluded, due to uninterpretable PCR results or LUS technical issues. Of the 134 remaining patients, 31 (23%) were classified as COVID^pos^ and 103 (77%) as COVID^neg^ based on Rt-PCR test. Among the 13 COVID^neg^ patients who had a second screening test during the 30-day follow-up, only one had a positive SARS-CoV-2 Rt-PCR, related to a clearly distinct clinical episode. This patient was thus classified as COVID^neg^. Most patients were female (63%), healthcare workers (85%) with a median age of 35 years; most sought out testing within the first 5 days of symptom onset (Table 1). COVID^pos^ patients had fewer comorbidities than COVID^neg^, the latter suffering mostly from asthma, obesity or hypertension. COVID^pos^ patients presented more often with a history of fever and anosmia, but less often with dyspnea than COVID^neg^ patients. Vital signs at inclusion were normal in most patients of both groups.

**Table 1.**
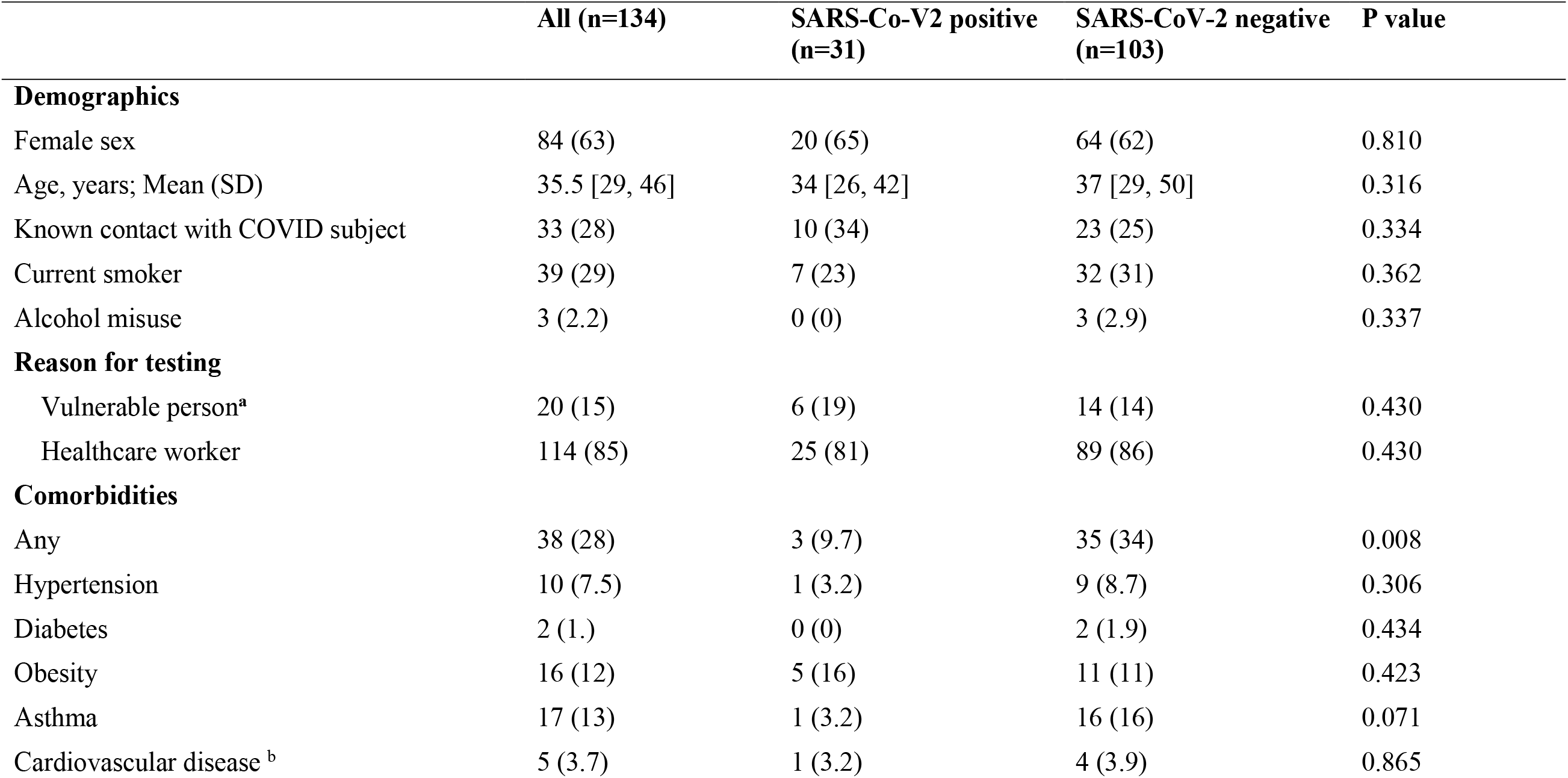
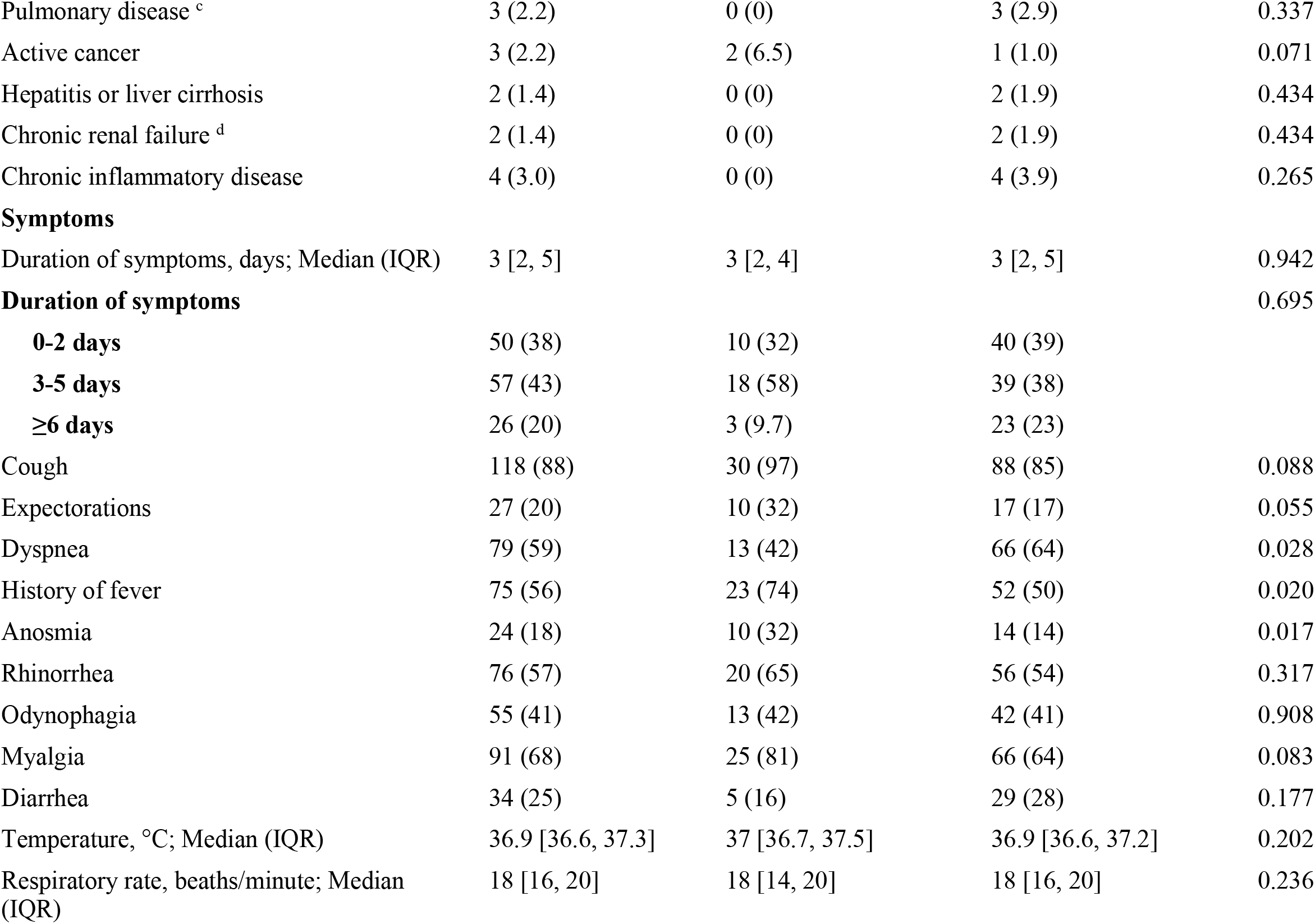
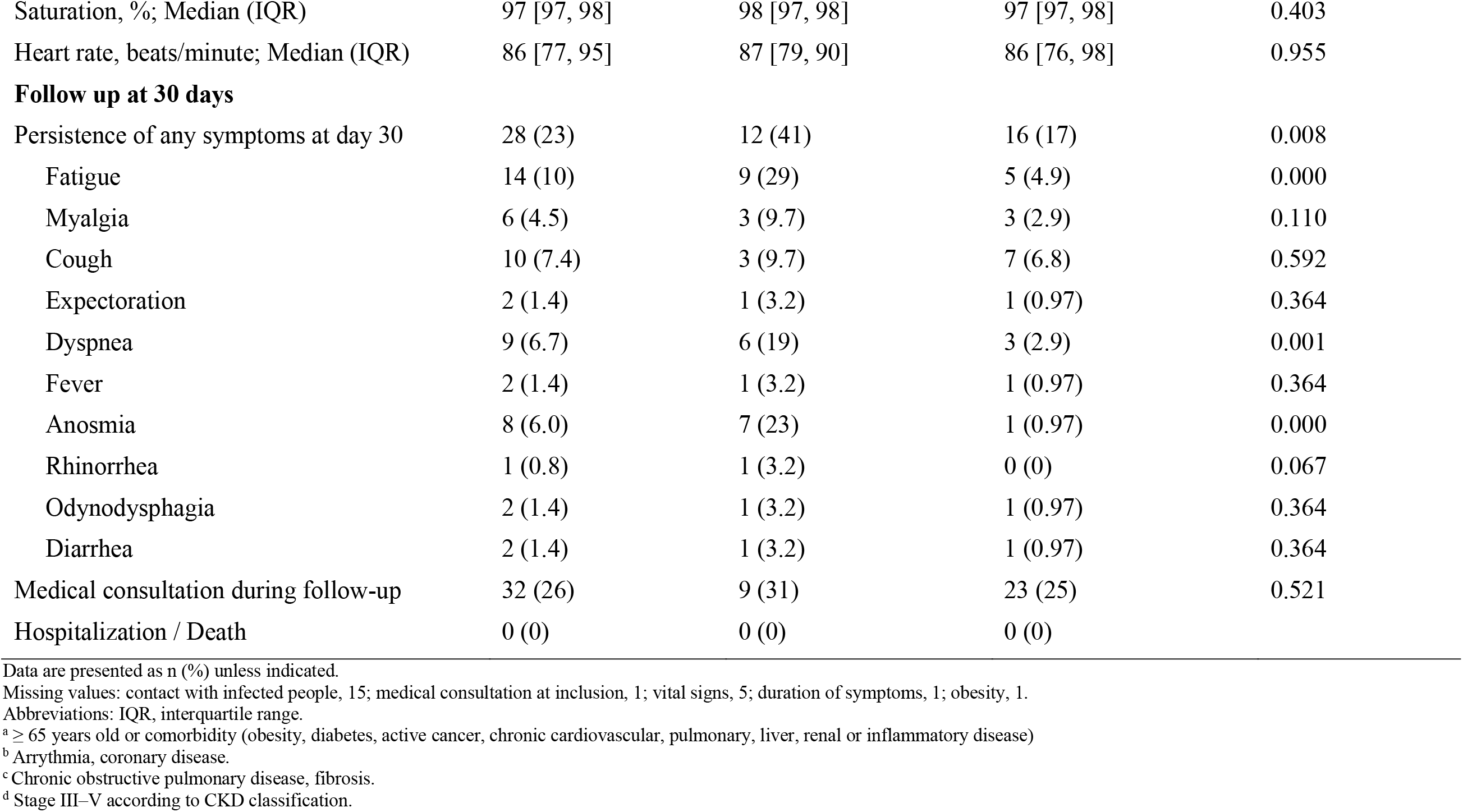
Demographics, clinical characteristics and 30-day outcome of study participants according to nasopharyngeal Rt-PCR SARS-CoV-2 results

|  | All (n=134) | SARS-Co-V2 positive (n=31) | SARS-CoV-2 negative (n=103) | P value |
| --- | --- | --- | --- | --- |
| **Demographics** |  |  |  |  |
| Female sex | 84 (63) | 20 (65) | 64 (62) | 0.810 |
| Age, years; Mean (SD) | 35.5 [29, 46] | 34 [26, 42] | 37 [29, 50] | 0.316 |
| Known contact with COVID subject | 33 (28) | 10 (34) | 23 (25) | 0.334 |
| Current smoker | 39 (29) | 7 (23) | 32 (31) | 0.362 |
| Alcohol misuse | 3 (2.2) | 0 (0) | 3 (2.9) | 0.337 |
| **Reason for testing** |  |  |  |  |
| Vulnerable person<sup>a</sup> | 20 (15) | 6 (19) | 14 (14) | 0.430 |
| Healthcare worker | 114 (85) | 25 (81) | 89 (86) | 0.430 |
| **Comorbidities** |  |  |  |  |
| Any | 38 (28) | 3 (9.7) | 35 (34) | 0.008 |
| Hypertension | 10 (7.5) | 1 (3.2) | 9 (8.7) | 0.306 |
| Diabetes | 2 (1.) | 0 (0) | 2 (1.9) | 0.434 |
| Obesity | 16 (12) | 5 (16) | 11 (11) | 0.423 |
| Asthma | 17 (13) | 1 (3.2) | 16 (16) | 0.071 |
| Cardiovascular disease <sup>b</sup> | 5 (3.7) | 1 (3.2) | 4 (3.9) | 0.865 |
| Pulmonary disease <sup>c</sup> | 3 (2.2) | 0 (0) | 3 (2.9) | 0.337 |
| Active cancer | 3 (2.2) | 2 (6.5) | 1 (1.0) | 0.071 |
| Hepatitis or liver cirrhosis | 2 (1.4) | 0 (0) | 2 (1.9) | 0.434 |
| Chronic renal failure <sup>d</sup> | 2 (1.4) | 0 (0) | 2 (1.9) | 0.434 |
| Chronic inflammatory disease | 4 (3.0) | 0 (0) | 4 (3.9) | 0.265 |
| **Symptoms** |  |  |  |  |
| Duration of symptoms, days; Median (IQR) | 3 [2, 5] | 3 [2, 4] | 3 [2, 5] | 0.942 |
| **Duration of symptoms** |  |  |  | 0.695 |
| **0-2 days** | 50 (38) | 10 (32) | 40 (39) |  |
| **3-5 days** | 57 (43) | 18 (58) | 39 (38) |  |
| **≥6 days** | 26 (20) | 3 (9.7) | 23 (23) |  |
| Cough | 118 (88) | 30 (97) | 88 (85) | 0.088 |
| Expectorations | 27 (20) | 10 (32) | 17 (17) | 0.055 |
| Dyspnea | 79 (59) | 13 (42) | 66 (64) | 0.028 |
| History of fever | 75 (56) | 23 (74) | 52 (50) | 0.020 |
| Anosmia | 24 (18) | 10 (32) | 14 (14) | 0.017 |
| Rhinorrhea | 76 (57) | 20 (65) | 56 (54) | 0.317 |
| Odynophagia | 55 (41) | 13 (42) | 42 (41) | 0.908 |
| Myalgia | 91 (68) | 25 (81) | 66 (64) | 0.083 |
| Diarrhea | 34 (25) | 5 (16) | 29 (28) | 0.177 |
| Temperature, °C; Median (IQR) | 36.9 [36.6, 37.3] | 37 [36.7, 37.5] | 36.9 [36.6, 37.2] | 0.202 |
| Respiratory rate, beaths/minute; Median (IQR) | 18 [16, 20] | 18 [14, 20] | 18 [16, 20] | 0.236 |
| Saturation, %; Median (IQR) | 97 [97, 98] | 98 [97, 98] | 97 [97, 98] | 0.403 |
| Heart rate, beats/minute; Median (IQR) | 86 [77, 95] | 87 [79, 90] | 86 [76, 98] | 0.955 |
| **Follow up at 30 days** |  |  |  |  |
| Persistence of any symptoms at day 30 | 28 (23) | 12 (41) | 16 (17) | 0.008 |
| Fatigue | 14 (10) | 9 (29) | 5 (4.9) | 0.000 |
| Myalgia | 6 (4.5) | 3 (9.7) | 3 (2.9) | 0.110 |
| Cough | 10 (7.4) | 3 (9.7) | 7 (6.8) | 0.592 |
| Expectoration | 2 (1.4) | 1 (3.2) | 1 (0.97) | 0.364 |
| Dyspnea | 9 (6.7) | 6 (19) | 3 (2.9) | 0.001 |
| Fever | 2 (1.4) | 1 (3.2) | 1 (0.97) | 0.364 |
| Anosmia | 8 (6.0) | 7 (23) | 1 (0.97) | 0.000 |
| Rhinorrhea | 1 (0.8) | 1 (3.2) | 0 (0) | 0.067 |
| Odynodysphagia | 2 (1.4) | 1 (3.2) | 1 (0.97) | 0.364 |
| Diarrhea | 2 (1.4) | 1 (3.2) | 1 (0.97) | 0.364 |
| Medical consultation during follow-up | 32 (26) | 9 (31) | 23 (25) | 0.521 |
| Hospitalization / Death | 0 (0) | 0 (0) | 0 (0) |  |

Data are presented as n (%) unless indicated.
Missing values: contact with infected people, 15; medical consultation at inclusion, 1; vital signs, 5; duration of symptoms, 1; obesity, 1.
Abbreviations: IQR, interquartile range.
<sup>a</sup> ≥ 65 years old or comorbidity (obesity, diabetes, active cancer, chronic cardiovascular, pulmonary, liver, renal or inflammatory disease)
<sup>b</sup> Arrhythmia, coronary disease.
<sup>c</sup> Chronic obstructive pulmonary disease, fibrosis.
<sup>d</sup> Stage III–V according to CKD classification.

### Lung ultrasonography findings

Lung ultrasound was abnormal in 31% of patients (Table 2). The two observers showed good concordance to differentiate a normal from an abnormal LUS, with a kappa of 0.67. Most anomalies were focal and unilateral. The most frequent patterns were pathologic B-lines and thickening of the pleura with pleural line irregularities. Only 9.1% of control subjects presented any abnormal finding on LUS, and all these anomalies were focal pathologic or confluent B lines (Supplementary Table 2).

**Table 2.**
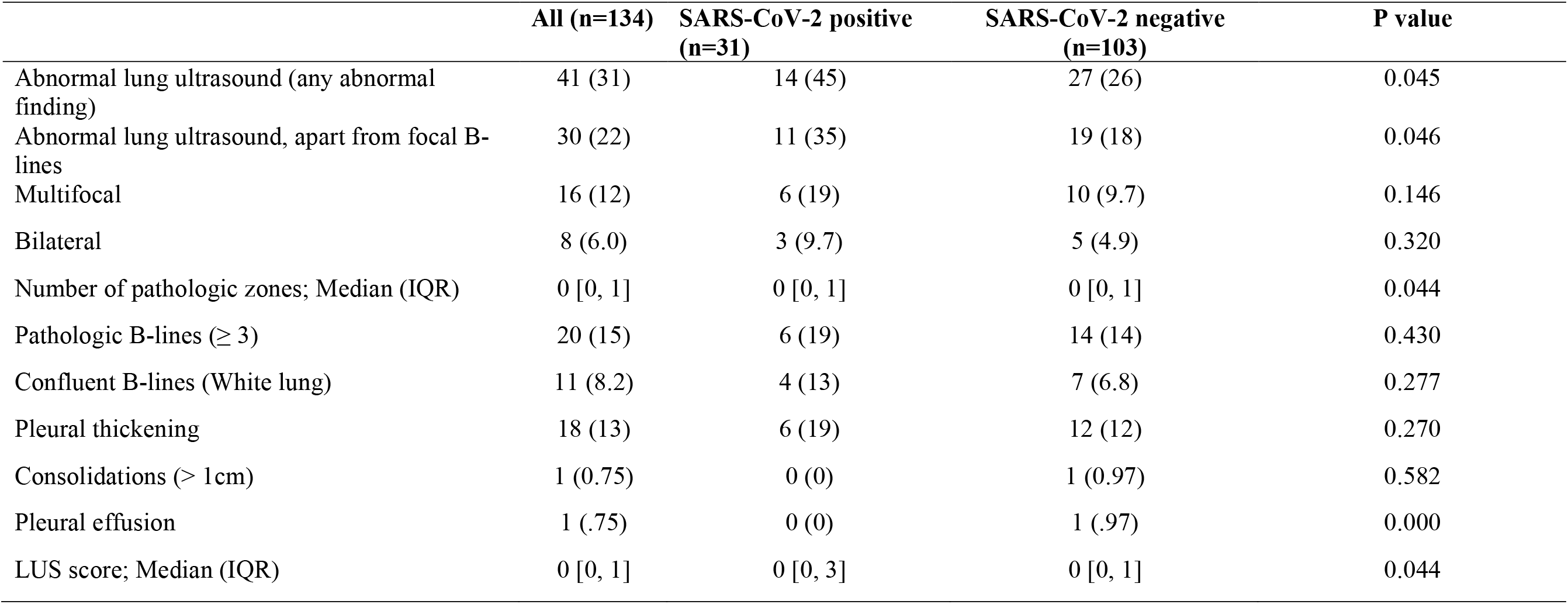
Lung ultrasound characteristics of study participants according to nasopharyngeal Rt-PCR SARS-CoV-2 results

Among all symptomatic patients, two factors were significantly associated with abnormal LUS: SARS-CoV-2 infection and history of fever (Table 3). Indeed, COVID^pos^ patients had abnormal LUS findings significantly more frequently compared with COVID^neg^ (45% versus 26%, p=0.045). However, this feature alone was poorly sensitive (45%) and specific (74%). No specific ultrasonographic pattern on its own significantly distinguished COVID^pos^ from COVID^neg^ subjects (Table 2).

**Table 3.** Demographics and clinical characteristics of study participants according to the presence of an abnormal lung ultrasound

|  | All (n=134) | Abnormal LUS (n=41) | Normal LUS (n=93) | P value |
| --- | --- | --- | --- | --- |
| **Demographics** |  |  |  |  |
| Female sex | 84 (63) | 28 (68) | 56 (60) | 0.373 |
| Age; Median (IQR) | 35.5 [29, 46] | 38 [31, 48] | 35 [28, 45] | 0.574 |
| Current cigarette smoker | 39 (29) | 12 (29) | 27 (29) | 0.978 |
| Alcohol misuse | 3 (2.2) | 0 (0) | 3 (3.2) | 0.245 |
| **Reason of testing** |  |  |  |  |
| Vulnerable person | 20 (15) | 3 (7.3) | 17 (18) | 0.101 |
| Healthcare worker | 114 (85) | 38 (93) | 76 (82) | 0.101 |
| **Positive Rt-PCR result** | 31 (23) | 14 (34) | 17 (18) | 0.045 |
| **Comorbidities** |  |  |  |  |
| Any | 38 (28) | 13 (32) | 25 (27) | 0.568 |
| Hypertension | 10 (7.5) | 3 (7.3) | 7 (7.5) | 0.966 |
| Diabetes | 2 (1.5) | 1 (2.4) | 1 (1.1) | 0.549 |
| Obesity | 16 (12) | 3 (7.3) | 13 (14) | 0.265 |
| Asthma | 17 (13) | 7 (17) | 10 (11) | 0.311 |
| Cardiovascular disease <sup>b</sup> | 5 (3.7) | 2 (4.9) | 3 (3.2) | 0.642 |
| Pulmonary disease <sup>c</sup> | 3 (2.2) | 0 (0) | 3 (3.2) | 0.245 |
| Active cancer | 3 (2.2) | 1 (2.4) | 2 (2.2) | 0.917 |
| Hepatitis or liver cirrhosis | 2 (1.5) | 1 (2.4) | 1 (1.1) | 0.549 |
| Chronic renal failure <sup>d</sup> | 2 (1.5) | 0 (0) | 2 (2.2) | 0.344 |
| Chronic inflammatory disease | 4 (3.0) | 0 (0) | 4 (4.3) | 0.178 |
| **Symptoms** |  |  |  |  |
| Duration of symptoms, days; Median (IQR) | 3 [2, 5] | 3 [2, 5] | 3 [2, 5] | 0.344 |
| Duration of symptoms |  |  |  | 0.210 |
| 0-2 days | 50 (38) | 11 (22) | 39 (78) |  |
| 3-5 days | 57 (43) | 21 (37) | 36 (63) |  |
| ≥ 6 days | 26 (20) | 9 (35) | 17 (65) |  |
| Cough | 118 (88) | 34 (83) | 84 (90) | 0.224 |
| Expectorations | 27 (20) | 7 (17) | 20 (22) | 0.556 |
| Dyspnea | 79 (59) | 25 (61) | 54 (58) | 0.752 |
| Hemoptysis | 2 (1.5) | 0 (0) | 2 (2.2) | 0.344 |
| History of fever | 75 (56) | 29 (71) | 46 (49) | 0.022 |
| Anosmia | 24 (18) | 11 (27) | 13 (14) | 0.074 |
| Rhinorrhea | 76 (57) | 21 (51) | 55 (59) | 0.394 |
| Odynophagia | 55 (41) | 17 (41) | 38 (41) | 0.948 |
| Myalgia | 91 (68) | 31 (76) | 60 (65) | 0.205 |
| Diarrhea | 34 (25) | 8 (20) | 26 (28) | 0.301 |
| Temperature, °C; Median (IQR) | 36.9 [36.6, 37.3] | 37 [36.6, 37.5] | 36.9 [36.6, 37.2] | 0.270 |
| Respiratory rate, breaths/minute; Median (IQR) | 18 [16, 20] | 18 [16, 20] | 18 [16, 20] | 0.330 |
| Saturation, %; Median (IQR) | 97 [97, 98] | 97 [97, 98] | 97 [97, 98] | 0.385 |
| Heart rate, beats/minute; Median (IQR) | 86 [77, 95] | 88 [79, 98] | 85 [76.5, 94] | 0.170 |
| **Follow-up at 30 days** |  |  |  |  |
| Persistence of any symptoms at day 30 | 28 (23) | 9 (24) | 19 (23) | 0.924 |
| Fatigue | 14 (10) | 7 (17) | 7 (7.5) | 0.096 |
| Myalgia | 6 (4.5) | 2 (4.9) | 4 (4.3) | 0.882 |
| Cough | 10 (7.5) | 3 (7.3) | 7 (7.5) | 0.966 |
| Expectorations | 2 (1.5) | 0 (0) | 2 (2.2) | 0.344 |
| Dyspnea | 9 (6.7) | 4 (9.8) | 5 (5.4) | 0.351 |
| Fever | 2 (1.5) | 0 (0) | 2 (2.2) | 0.344 |
| Anosmia | 8 (6.0) | 1 (2.4) | 7 (7.5) | 0.252 |
| Rhinorrhea | 1 (.75) | 0 (0) | 1 (1.1) | 0.505 |
| Odynophagia | 2 (1.5) | 1 (2.4) | 1 (1.1) | 0.549 |
| Diarrhea | 2 (1.5) | 0 (0) | 2 (2.2) | 0.344 |
| Medical consultation during follow-up | 26 (21) | 10 (26) | 16 (19) | 0.364 |
| Hospitalization/Death | 0 (0) | 0 (0) | 0 (0) |  |

Data are presented as n (%) unless otherwise indicated.
Abbreviations: IQR, interquartile range.
<sup>a</sup> ≥ 65 years old or comorbidity (obesity, diabetes, active cancer, chronic cardiovascular, pulmonary, liver, renal or inflammatory disease)
<sup>b</sup> Arrhythmia, coronary disease.
<sup>c</sup> Chronic obstructive pulmonary disease, fibrosis.
<sup>d</sup> Stage III–V according to CKD classification

Although not statistically different, the proportion of COVID-19^pos^ with abnormal LUS findings was positively associated with symptoms duration. While only 30% of COVID-19^pos^ patients had abnormal LUS within 2 days of symptom onset, 52% of patients had pathological LUS after 2 days (p=0.24).

### Multivariate diagnostic score

We combined LUS findings with symptoms, vital signs and a binary feature for known contact with a COVID-19 case to build a multivariate logistic regression diagnostic score. Using all features, the score had 78.8% sensitivity, 84.0% specificity, 83.1% PPV, 61.4% NPV, 4.9 LR+, 0.3 LR- and 84.5% AUC (Figure 1). We present a plot on which to assess the score according to a desired sensitivity/specificity trade-off.

**Figure 1.**
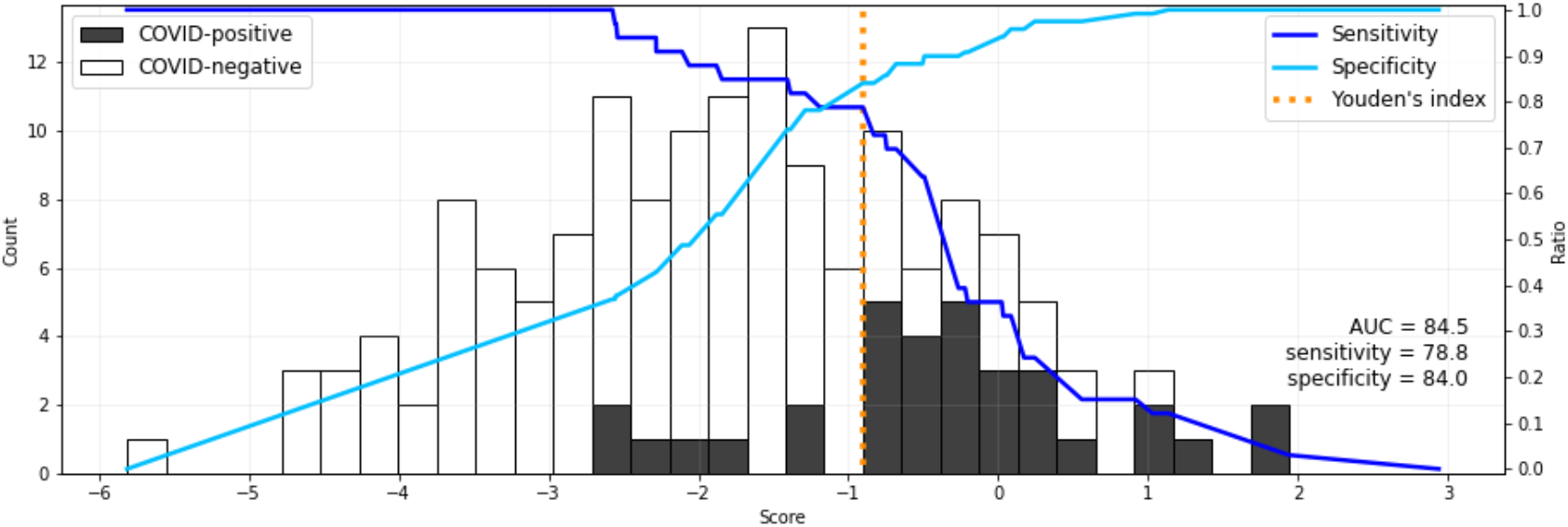
A multivariate logistic regression diagnostic score (x-axis) to discriminate COVID^pos^ from COVID^neg^ patients (black and white bars respectively with count on y axis). Sensitivity **(—)** and specificity **(—)** of the score are plotted with Youden’s index (sensitivity + specificity −1) marked in orange. All 22 features are used in the depicted image on a model trained on all data points.

In Table 4, score performance with several combinations of features at various stages of RFE are presented. The strongest positive predictor was any evidence of pleural thickening at any number of sites (coefficient: +0.69) with LUS, although it became a negative predictor with an increasing number of sites with this feature (−0.40). The presence of pathological B lines and confluent pathological B lines were also positively associated with COVID infection in this score. All three of the above patterns were retained by RFE within the top seven features. The LUS features that were negative and quickly eliminated by RFE were those describing consolidation and multifocal pathology. Cough, fever and anosmia were the highest ranked symptoms (coefficient ≥0.4), in line with previous reports. While LUS patterns were highly ranked in the RFE, rerunning the model without LUS findings reduced AUC by only 4% (AUC 84.5% vs 80.3%). LUS findings were poorly sensitive in the absence of clinical features (AUC: 63.9% Sensitivity: 45.5%, Specificity: 77.3%, PPV: 66.7%, NPV: 55.6%, LR+: 2.0, LR-: 0.7).

**Table 4.**
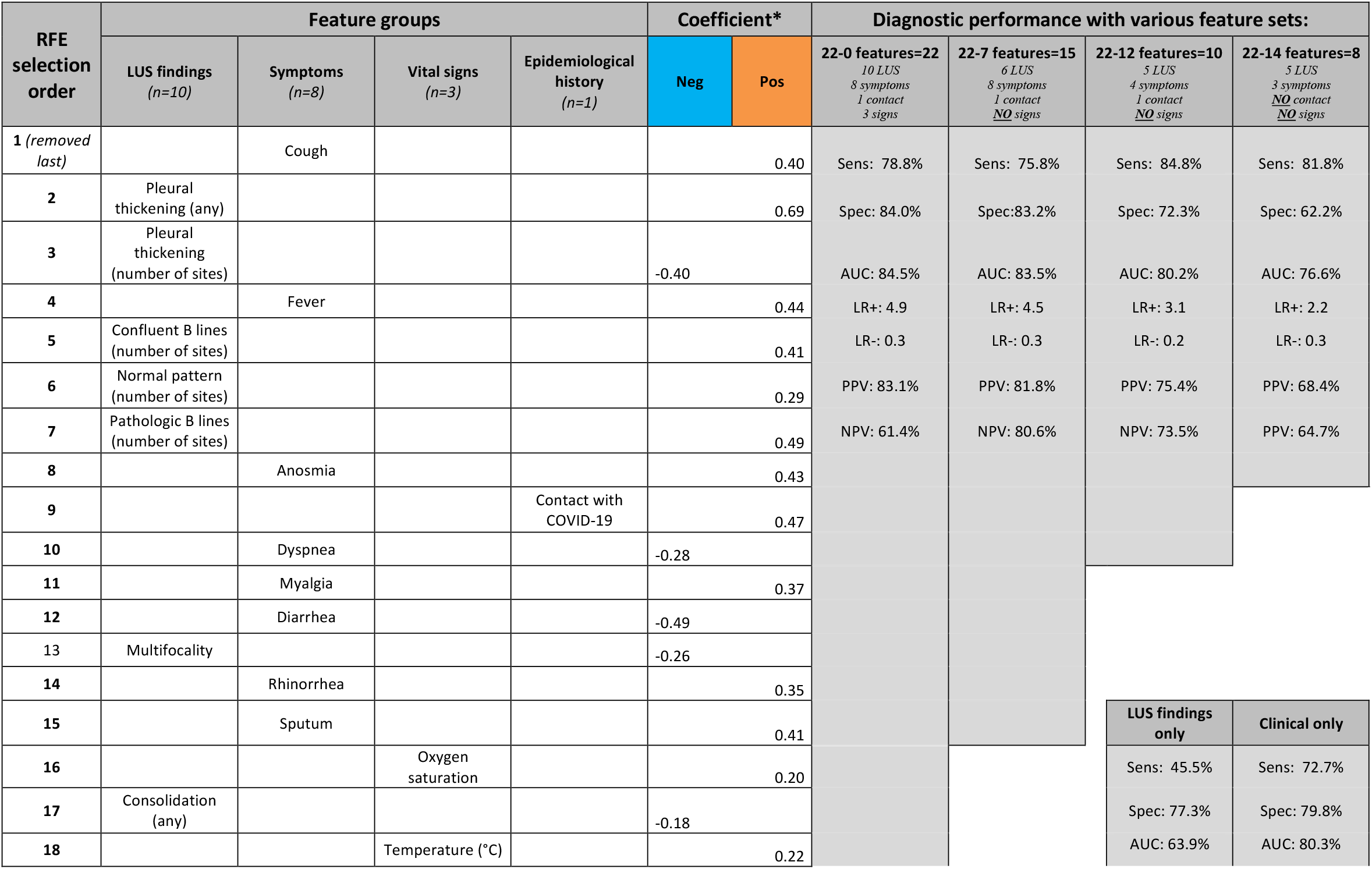
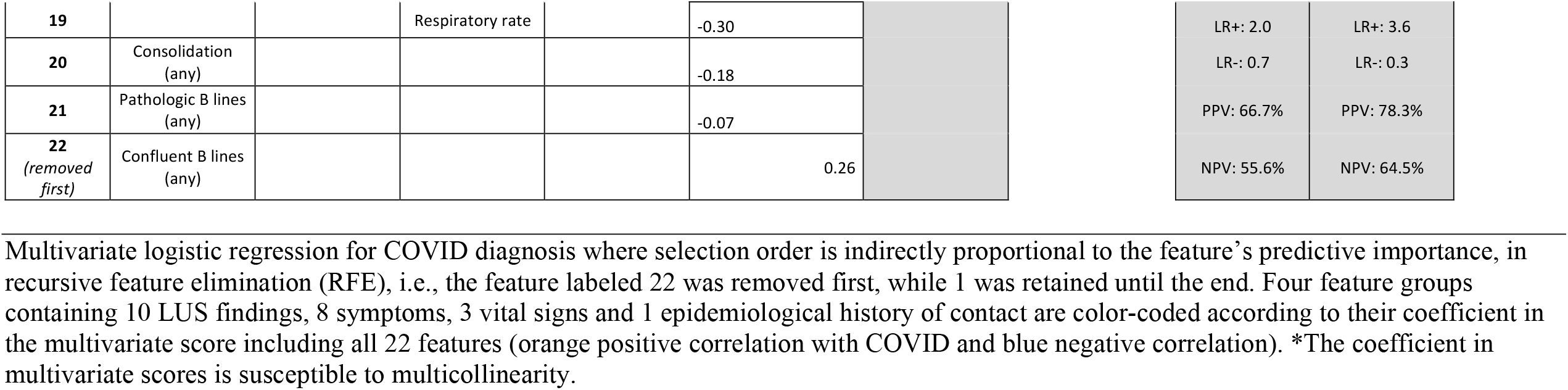
Multivariate logistic regression for COVID diagnosis

Combining all 22 features and using RFE, we observe that removing 7 features had minimal impact on score performance, and removing 12 features reduces AUC by only 4% compared to the original.

### 30-day outcome

The 30-day follow-up was available for 121/134 (90%) patients. None was hospitalized or died during follow-up. COVID^pos^ patients had more frequently persistent symptoms (fatigue, dyspnea or anosmia) at 30-day compared with COVID^neg^ (Table 1).

The presence of an abnormal LUS at inclusion was not associated with symptom persistence (Table 3).

As no patients were admitted or died, we could not analyze the value of LUS findings to predict critical clinical outcome.

## Discussion

Lung pathology is detectable by chest CT early in the course of COVID disease, even in asymptomatic patients, suggesting that lung imaging might have a place as a complementary diagnostic tool [3]. However, large scale CT screening is not feasible even in hospital settings with abundant resources. Point-of-care LUS is now affordable, portable and implementable in a decentralized setting and has all the attributes to become a pragmatic community-based screening tool.

We evaluated the diagnostic performance of LUS in a prospective cohort of subjects with mild acute respiratory tract infection attending a COVID-19 Swiss screening center. COVID^pos^ outpatients more frequently had abnormal LUS findings at inclusion compared with COVID^neg^. However, LUS findings alone had insufficient sensitivity, NPV and LR-to recommend LUS as an independent screening tool in outpatients. The combination of LUS findings with clinical presentation showed promising results.

The limited sensitivity of LUS in our population is discordant with previous studies, which showed a good sensitivity (89-97%) to identify Rt-PCR-confirmed COVID-19. These retrospective studies were conducted in emergency departments and included patients with severe and critical COVID-19 infection[17–19]. Other studies using chest CT also showed an excellent sensitivity (97-98%) to diagnose COVID-19 [2,20,21]. However, all these studies were conducted in hospitalized patients with severe or critical disease, preventing extrapolation to our milder population screened for symptoms only.

The clinical severity of the disease strongly affects the performance of diagnostic tests, and particularly the sensitivity of LUS. We conclude that while LUS may be an interesting COVID-19 screening tool in emergency departments, it is not reliable when used alone in patients with mild disease. In the only study investigating chest CT features in patients with asymptomatic (73%) or mild (27%) COVID-19, which was conducted in the passengers of the cruise ship *Diamond Princess*, 54% of asymptomatic patients and 79% of patients with mild disease presented opacities on chest CT. These results suggested the potential use of chest CT in clinical decision making [3]. Most opacities were located in the peripheral areas of the lung, where LUS is performant. Patients included in the *Diamond Princess* study were older compared with our study population (mean of 63 ± 15 years vs. 39 ± 13 years), a possible explanation for the lower proportion of patients with lung involvement in our study. However, our data suggest that a combination of LUS findings and clinical characteristics might achieve better detection of mild COVID-19 in young outpatients.

We observed more abnormal LUS findings in COVID^pos^ patients who had more than 2 days of symptoms (52% versus 30%), although our results were not statistically significant. Concordant with our findings, a relationship between the duration of infection and the proportion of abnormal radiological findings has been described [22–24]. In one study, only 44% of patients presenting within 2 days of symptoms had an abnormal CT, while this proportion rose to 91% after 3 to 5 days and 96% after 5 days [24]. This study did not provide any data on COVID-19 severity. In another study using chest X-ray in patients admitted to the emergency department, the proportion of an abnormal chest X ray increased with the duration of symptoms (63% in the first 2 days to 84% after 9 days) [25].

In our study, most patients with abnormal LUS findings presented with focal pathologic B lines, confluent B lines or pleural thickening, irrespective of the etiology of the acute respiratory tract infection. Inclusion of healthy volunteers confirmed the causality between LUS findings and acute respiratory tract infections. Indeed, only 9% of healthy volunteers presented LUS anomalies (and all were focal pathologic B lines).

Two previous study showed that thickened pleural lines on LUS were significantly associated with COVID-19 [17,18]. However, in a third report, LUS findings were similar in both COVID-19 and non-COVID-19 patients [19].

### Limitations

Our study has some limitations. First, most of our subjects were healthy and young healthcare workers, which prevents extrapolation of our results to an older and comorbid population. However, young, healthy subjects are of a prime importance in the management of the virus spread [26]. Second, SARS-CoV-2 Rt-PCR nasopharyngeal swab was used as the gold standard, and we might have missed some early infections when it has limited sensitivity [27]. However, it is considered as the reference diagnostic method. Furthermore, we sought to mitigate technical and sample collection error using validated nucleic acid amplification tests and a dedicated trained medical team performing nasopharyngeal swabs [28]. In addition, we had 30-day follow-up, which may have reduced the number of patients misclassified as COVID^neg^. To better investigate the predictive potential of LUS findings, we built a multivariate score. The small sample size and high feature count (n= 22) exposes the model to the risk of overfitting. Thus, this score is not ready for clinical use, but rather is a mean to demonstrate the feature importance by RFE.

## Conclusion

To our knowledge, this is the first study, which assessed the use of LUS in a screening center outpatient population with mild COVID-19. As disease severity plays an important role in the ultrasonographic findings, LUS is poorly sensitive as a SARS-CoV-2 screening tool in the context of mild community-level screening. However, the good performance of a combination of clinical and LUS features showed promising results, which could be used to avoid a PCR test in patients with a negative screening. These results need confirmation in a larger study population.

## Data Availability

All data files are available at https://zenodo.org/record/4617904#.YFmlHi3pNQI

## Declarations

## Funding

This work was supported by an academic award of the Leenaards Foundation (to NBB), by the Foundation of Lausanne University Hospital, and the Emergency Department Lausanne University Hospital. The funding bodies had no role in the design of the study and collection, analysis and interpretation of data and in writing the manuscript.

## Acknowledgements

We thank all the patients who accepted to participate and make this study possible. We thank all healthcare workers of the triage unit of the emergency department of the University Hospital of Lausanne, who supported the study and managed COVID-19 suspected patients.

## Authors’ contributions

JYM, OH, PC, NBB: study conception, study design, study performance, study management, data analysis, data interpretation and manuscript writing. SS, TB, JYM, OP: LUS images review, data interpretation and critical review of the manuscript.

TE, LB: LUS images recording, data interpretation and critical review of the manuscript. MH, JC: data analysis, interpretation, visualisations and critical review of the manuscript.

All authors approved the final version of the manuscript and agreed to be accountable for all aspects of the work in ensuring that questions related to the accuracy or integrity of any part of the work are appropriately investigated and resolved.

NBB had full access to all the data in the study and takes responsibility for the integrity of the data and the accuracy of the data analysis.

## List of Supplemental Digital Content

SupplementaryTables.docx

## Notes

**Funding and support:** this work was supported by the Leenards foundation and by Lausanne University Hospital

### Competing Interest Statement

The authors have declared no competing interest.

### Author Declarations

The study was approved by the Swiss Ethics Committee of the canton of Vaud (CER-VD 2019-02283).

