## Supplementary Tables for "Point-of-care lung ultrasonography for early identification of mild COVID-19: a prospective cohort of outpatients in a Swiss screening center"

**Supplementary Table 1.** Characteristics of study participants comparing healthy controls and patients with a lower respiratory tract infection (COVID^pos^ and COVID^neg^).

|  | **All (n=178)** | **LRTI patients (n=134)** | **Control patients (n=44)** | **P value** |
| --- | --- | --- | --- | --- |
| Female sex | 112 (63) | 84 (63) | 28 (64) | 0.910 |
| Age, years; Median (IQR) | 34 [28, 45] | 35 [29, 46] | 31 [25, 42] * | 0.007 |
| Pulmonary disease ^a^ | 3 (1.7) | 3 (2.2) | 0 (0) | 0.317 |
| Current cigarettes smoker | 51 (29) | 39 (29) | 12 (27) | 0.816 |

*** *p* < 0.05

Data are presented as n (%) unless otherwise indicated.

Missing values: 0

Abbreviations: IQR, interquartile range; LRTI, Lower respiratory tract infection

^a^ COPD, fibrosis.

**Supplementary Table 2.** Lung ultrasound characteristics of study participants comparing healthy controls and patients with a lower respiratory tract infection (COVID^pos^ and COVID^neg^).

|  | **All**  **(n=178)** | **LRTI patients**  **(n=134)** | **Control patients**  **(n=44)** | **P value** |
| --- | --- | --- | --- | --- |
| Abnormal lung ultrasound | 45 (25) | 41 (31) | 4 (9.1) * | 0.004 |
| Abnormal lung ultrasound apart from focal B lines | 31 (17) | 30 (22) | 1 (2.2) | 0.002 |
| Multifocal | 16 (9.0) | 16 (12) | 0 (0) * | 0.016 |
| Bilateral | 8 (4.5) | 8 (6.0) | 0 (0) | 0.097 |
| Number of pathologic zones; Median (IQR) | 0 [0, 0.7] | 0 [0, 1] | 0 [0, 0] * | 0.003 |
| Pathologic B lines (≥3) | 23 (13) | 20 (15) | 3 (6.8) | 0.164 |
| Confluent B lines (White lung) | 12 (6.7) | 11 (8.2) | 1 (2.3) | 0.173 |
| Thickening of the pleura with pleural line irregularities | 18 (10) | 18 (13) | 0 (0) * | 0.010 |
| Consolidations (>1cm) | 1 (0.6) | 1 (0.8) | 0 (0) | 0.566 |
| Pleural effusion | 0 (0) | 0 (0) | 0 (0) |  |
| LUS score; Median (IQR) | 0 [0, 0.75] | 0 [0, 1] | 0 [0, 0] * | 0.003 |

Data are presented as n (%) unless otherwise indicated.

Abbreviations: IQR, interquartile range.
